# Application of aptamer technology in enterobacteria and non-fermenters: literature review

**DOI:** 10.1101/2022.05.11.22274950

**Authors:** Taniela Marli Bes, Marina Cortês Farrel, Carlos Santos, Ester Cerdeira Sabino, Silvia Figueiredo Costa

## Abstract

Antimicrobial resistance has grown exponentially in the last decade and become a global health threat. The antibiotic resistance crisis has guided the scientific community to explore non-conventional interventions to target resistant bacteria. Development of new technologies, such as aptamers-based treatment and diagnosis, has shown to be promising with remarkable advantages over the past five years. This narrative review aims on what is already known regarding application of aptamer technology in enterobacteria and non-fermenters, and the prospects for future achievements. A systematic search of the English literature was performed on the 7th of December 2021 to identify papers on aptamer discovery, with a focus on gram negative isolates, published from January 01, 1993, to December 07, 2021, under the topics: (aptamer OR aptamers OR SELEX) AND (bacteria OR sepsis OR non-fermenter OR Enterobacteriaceae OR infection)). The reference lists of included articles were also searched, in addition to hand-searching of various relevant high-impact journals. Out of 2,474 articles, 30 experimental studies were recruited for review, and are chronologically described. Although the number of publications regarding development of aptamers to target these pathogenic agents has increased over the years, the recent publications are mostly around diagnostic devices manufactured using previously described aptamers. There have been less than one-third of the studies describing new and specific aptamers. From the 30 selected papers, 18 are regarding non-fermenters, seven approaching multi-species of bacteria and only five regarding a single enterobacteria. Even for the newly described aptamers, most of the published papers pertain to diagnostic aptamers and only seven focus on aptamers for therapeutics. The number of aptamers with strong and specific binding capacity are still limited. Improving the current SELEX and developing more APT remains the major hurdle for aptamer related studies.

## Introduction

In the era of DNA-based therapy, bacterial infections still struggle with only a few therapeutic options. Currently, the main concern regarding bacterial infections is focused on the Gram-negative bacteria. Although the Enterobacteriaceae family by itself includes more than 70 genera, all the attention is towards *Klebsiella spp*., *Enterobacter spp*., *and the non-fermenters Acinetobacter spp*. and *Pseudomonas spp*., the most common agents involved in health-care– acquired infections. They are responsible for thousands of deaths every year, associated with millions of dollars in healthcare costs (CDC, 2019). The Gram-negative bacteria brings an extra challenge; besides the usual antibiotic resistance mechanisms, their outer membrane evolves and adapts to the protective mechanisms against the selection pressure of antibiotics [1].

Antimicrobial resistance has grown exponentially in the last decade and become a global health threat. Regardless of the emergence of multidrug-resistant (MDR) microorganisms in the community and healthcare setting, the treatment options are limited. Since April of 2020, only 41 new antibiotics have been in development and, out of those, only 13 have potential activity against carbapenem-resistant/extended spectrum β-lactamase (ESBL)-producing Enterobacteriaceae (pewtrusts.org).

The antibiotic resistance crisis has guided the scientific community to think about non-conventional interventions to limit the emergence and spread of MDR bacteria. Development of new technologies, such as aptamer-based treatment and diagnosis, has been promising. The aptamer technology, first described in 1990, is an innovative tool based on RNA, modified RNA, single-stranded DNA (ssDNA), or double-stranded DNA oligonucleotides with special folding and bonding ability [2]. Thus far, aptamer targets can be varied from simple ions, low-molecular-weight ligands, and proteins to whole cells [3,4].

Due to numerous advantages, such as stability, range of targets, and diverse applications, aptamers are considered a therapeutic beyond the scope of the existing pharmacological drugs. They are generated by a method called SELEX (Systematic Evolution of Ligands via Exponential Enrichment), a time and labor-consuming protocol that has been optimized over the past thirty years with a common goal of producing novel aptamers for specific targets [5,6]. The concept of SELEX relies on the ability of aptamers to fold into unique three-dimensional structures in the presence of their respective targets, and bind with high affinity and specificity, being able to discriminate closely-related ligands (Figure 1).

**Figure 1:**
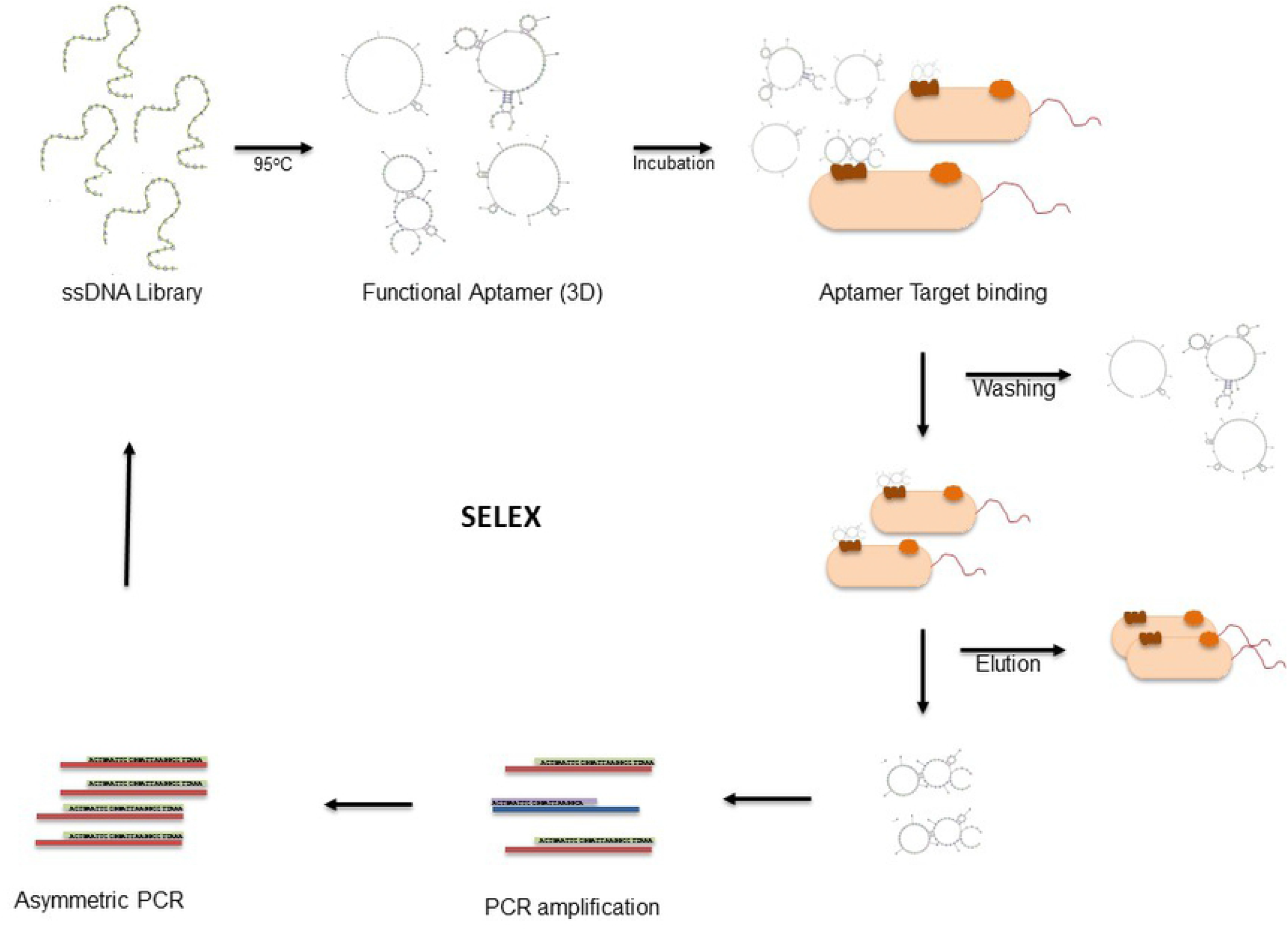
Selex protocol

The SELEX techniques have been modified to allow aptamer selection against living bacteria cells [4,7,8]. The so-called Cell-SELEX can be used to select aptamers against bacteria cell with no previous knowledge of the binding target and approaching the pathogen close to their native conformations and physiological environment.

Besides of innumerous hurdles in aptamer development, aptamer-based studies have shown remarkable advantages over the past five years. Unfortunately, there are limited options regarding gram negative bacteria so far.

The attractive future of aptamer technology, added to the clinical and epidemiologic concerns on antimicrobial resistance, has raised the possibility of using aptamer-based therapeutics and diagnostic tool for bacterial infections. This narrative review focuses on what we previously known regarding the application of aptamer technology for the more concerning Gram-negative bacteria and the prospects for future achievement.

## Methodology

### Search strategy

A systematic search of the English literature was performed on December 07, 2021, to identify papers on aptamer discovery, focused on gram negative isolates, published from January 01, 1993, to December 07, 2021. The date was selected because the original research on aptamer discovery was from this date. An initial electronic search was conducted using PubMed database, looking for original articles concerning “What is already established regarding aptamer-based technology for gram negative bacterial cells?” by manually searching all articles within the studied period under the topics: (aptamer OR aptamers OR SELEX) AND (bacteria OR sepsis OR non-fermenter OR Enterobacteriaceae OR *Klebsiella pneumoniae* OR *Acinetobacter baumannii* OR *Pseudomonas aeruginosa* OR infection). The reference lists of included articles were also searched, in addition to hand-searching of various relevant high-impact journals.

We used Mendeley desktop Software to check for any duplicate and allocate each reference to two independent reviewers, who screened it for inclusion based on its title and abstract (T.B. and M.F.). In case of doubts, the mentor (S.C.) was consulted. Full-text copies of all publications eligible for inclusion were subsequently assessed by 2 independent reviewers (T.B. and M.F.) and included when they met our prespecified inclusion criteria. Disagreement was solved through discussion or by consulting a third investigator (S.C.). The search strategy is detailed in the flow diagram in Figure 2. Out of 2,474 articles, 30 experimental studies were recruited for review.

**Figure 2:**
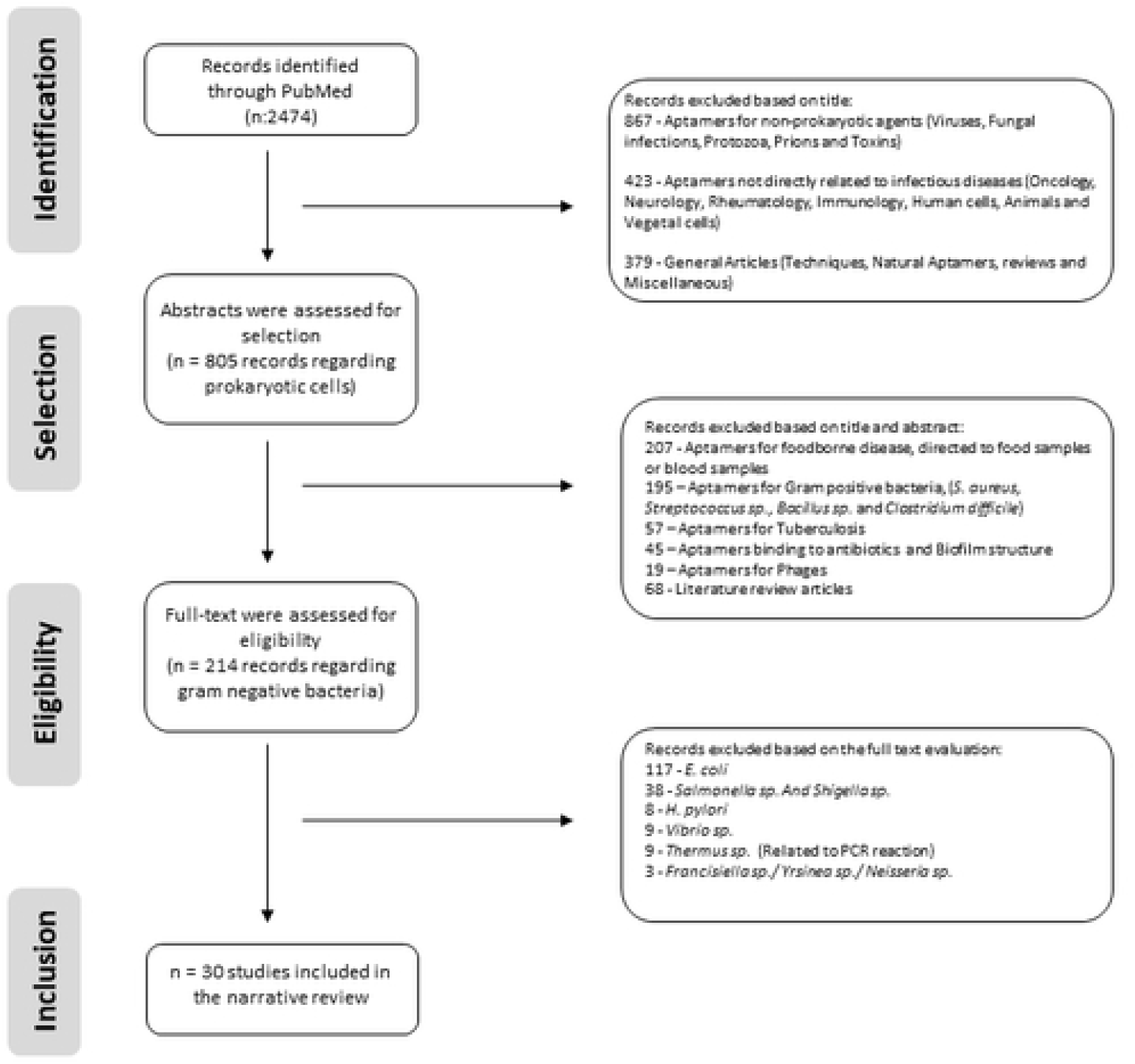
Flowchart of selected studies

### Study characteristics and data extraction

From the included studies, we registered bibliographic data such as title, authors, year of publication, journal of publication, and language. We also extracted data on study design, such as aptamer targets, type of sample, type of selection protocol, the aptamer function (therapy, diagnostic, or drug delivery). When there were two or more replications of the same experiment, they were analyzed separately and described in the discussion. If the outcome measured data were missing, we attempted to contact the authors for additional information. If the data could not be obtained, when possible, a conservative estimate was used, otherwise, a blank space was left.

**Table 1:** Aptamers used in detection of bacteria and their products. Table 1 shows in detail the 30 records used for this review. APT: Aptamer; CPX: Ciprofloxacin; FAM: Fluorescein amidite; Kd: Dissociation constant; LPS: lipopolysaccharide; NECEEM: Nonequilibrium capillary electrophoresis of equilibrium mixtures; QD: quantum dots; RNA: Ribonucleic acid; SELEX: Systematic Evolution of Ligands via Exponential Enrichment; ssDNA: Single-Stranded Deoxyribonucleic acid; SiC: Silicon carbide; SWNTs: Single-walled carbon nanotubes; STC-SELEX: sequential toggle cell-SELEX

| Reference<br>Year of<br>publication | Title | Microorganis<br>m | Technique | Specificity/Affinity | Goal | Specific target |
| --- | --- | --- | --- | --- | --- | --- |
| Ochsner UA and Vasil ML, 1996 | Gene repression by the ferric uptake regulator in *P. aeruginosa*: Cycle selection of iron-regulated genes | *P. aeruginosa* | SELEX (RNA) | DNA mobility-shift assays: the clones contained a Fur-box sequence identity | Gene identification | Identification of 16 novel genes controlled by the Fur repressor |
| Gnanam et al, 2008 | Development of aptamers specific for potential diagnostic targets in *Burkholderia pseudomallei*. | *Burkholderia pseudomallei* | Protein SELEX (RNA) | After the second round, negative selections were performed by passing the RNA over filters before protein incubation. | Diagnostic | *B. pseudomallei* proteins:<br>BipD<br>BopE<br>BPSL2748 |
| Ding JL, Gan ST and Ho B, 2009 | Single-Stranded DNA Oligoaptamers: Molecular Recognition and Lipopolysaccharide (LPS) Antagonism Are Length and Secondary Structure-Dependent. | Multiple | SELEX (DNA) | Tested specificity through sequential rounds of SELEX (7) followed by analyses of the bp sequencing and affinity of interaction between the oligoaptamers using the Kd values | Sepsis Treatment | ssDNA that binds LPS/Lipid A specifically. |
| Kim et al,2012 | Harnessing aptamers for electrochemical detection of endotoxin | Multiple | Non-SELEX (Nonequilibrium capillary electrophoresis of equilibrium mixtures (NECEEM) (DNA) | Specificity was tested doing negative selection with an APT sequence without affinity to LPS. Affinity was determined through the Kd using electropherogram. | LPS identification | LPS |
| Wang et al, 2011 | Utility of aptamer-fluorescence in situ hybridization for rapid detection of *P. aeruginosa*. | *P. aeruginosa* | Whole bacterium SELEX (DNA) | The binding affinity of ssDNA aptamers was determined by performing flow-cytometric analysis with increasing amount of the aptamer clone and a constant amount of *P. aeruginosa* (107) for each assay. For the specificity test, the flow-cytometry was performed with *P. aeruginosa* and other *Pseudomonas* spp., as well as tested against other gram-negative and gram-positive bacterial species. | Diagnostic<br>F17<br>F23 | Whole bacterial cell surface |
| Savory et al, 2015 | In silico maturation of binding-specificity of | *Proteus mirabilis* | Cell-SELEX | Affinity was assessed using a blotting assay in which *P.* | Diagnostic | Whole bacterial cell surface |
|  | DNA aptamers against *Proteus mirabilis*. |  | (DNA) | *mirabilis* cells immobilized on a nitrocellulose membrane were incubated with a range of concentrations of the APTs and the Kd was accessed.<br>Specificity: bacteria from other species and others genus were immobilized on a single piece of nitrocellulose membrane and incubated with labeled aptamer. | PmA102<br>PmA109 |  |
| Yoo SM, Kim D-K and Lee SY, 2015 | Aptamer-functionalized localized surface plasmon resonance sensor for the multiplexed detection of different bacterial species. | *Lactobacillus acidophilus*<br>*Salmonella typhimurium*<br>*Pseudomonas aeruginosa* | Non-SELEX<br>(DNA) | Utilized a modified Savory et al APT | Diagnostic | Whole bacterial cells |
| Rasoulinejad S and Gargari SLM, 2016 | Aptamer-nanobody based ELASA for specific detection of *Acinetobacter baumannii* isolates. | *A. baumannii* | Whole bacterium SELEX<br>(DNA) | Specificity was measured by flow cytometry after incubation with bacteria from another genus.<br>Affinity: Kd was measured after incubation with varying concentrations of aptamers | Diagnostic<br>Aci49<br>Amino modified aptamers connected to a silicon carbide (SiC) quantum dots (QDs) surface, resulting in a DNA-SiC QDs aptasensor for bacteria detection | Whole bacterial cells |
| Schulmeyer et al, 2016 | Primary and Secondary Sequence Structure Requirements for Recognition and Discrimination of Target RNAs by *Pseudomonas aeruginosa* RsmA and RsmF. | *P. aeruginosa* | SELEX<br>(RNA) | The radiolabeled RNA pool was submitted to affinity chromatography analyses and electrophoretic mobility shift assay. The aptamers equilibrium binding constant (K<sub>eq</sub>) was calculated. | Prediction of in vivo RNA targets of RsmA and RsmF | RsmA and RsmF |
| Soundy J and Day D, 2017 | Selection of DNA aptamers specific for live *Pseudomonas aeruginosa*. | *P. aeruginosa* | Whole bacterium SELEX (DNA) | Affinity: Flow cytometric analysis of fluorescently labelled aptamer pools. The dissociation constants were determined incubating bacterial cells with aptamer candidate concentrations in different ranges.<br>Specificity: aptamers were evaluated for binding to other Gram-negative bacterial species and Gram-positive. | Therapeutic | Against live biofilm derived from PA692 *P. aeruginosa* |
| Song et al, 2017 | Broadly reactive aptamers targeting bacteria belonging to different genera using a sequential toggle cell-SELEX. | *Escherichia coli*<br>*Enterobacter aerogenes*<br>*K. pneumoniae*<br>*Citrobacter freundii*<br>*Bacillus subtilis*<br>*Staphylococcus epidermidis* | STC-SELEX Process (DNA) | Affinity and specificity were investigated by binding aptamers to six target bacterial cells individually. For the selectivity test, the binding of the individual ssDNA sequences to different bacterial cells was performed using the same conditions. The fluorescence intensity of cells bound to 3' -FAM dye-labelled aptamers was measured using a micro-fluorospectrophotometer. The K<sub>d</sub> was obtained by a non-linear regression analysis | Diagnosis and treatment | Whole bacterial cells |
| Graziani et al, 2017 | High Efficiency Binding Aptamers for a Wide Range of Bacterial Sepsis Agents. | *A. baumannii*<br>*Listeria monocytogenes*<br>*E. coli*<br>*Staphylococcus aureus*<br>*K. pneumoniae* | SELEX (DNA) | Using real-time quantitative PCR, they access binding affinity comparing the Antibac1 and Antibac2 with a random library and through the dissociation constant (K<sub>d</sub>) | Antibac1 and Antibac2 - multiplex detection of bacterial sepsis in the clinical setting | Outer cell wall peptidoglycan |
| Wu et al, 2018 | A nitrocellulose membrane-based integrated microfluidic system for bacterial detection utilizing magnetic-composite membrane microdevices and | *A. baumannii* | SELEX (DNA) | The aptamer (10 μM) was load onto a NC membrane and immobilized via UV cross-linking. Then, 10<sup>3</sup> CFU/μL of *A. baumannii* (AB), *E. coli* (EC), and MRSA were incubated with the AB-specific aptamer and 30 min later the AB-specific, | Diagnostic | Whole bacterial cells |
|  | bacteria-specific aptamers. |  |  | biotin-conjugated aptamer was added to capture the membrane-bound bacteria. The aptamer was deemed to be AB-specific. |  |  |
| Wang et al, 2018 | Influence of aptamer-targeted antibiofilm agents for treatment of *Pseudomonas aeruginosa* biofilms. | *P. aeruginosa* | Non-SELEX<br>Aptamer-CPX-SWNTs (DNA) | Utilized a modified Wang et al APT | Biofilm treatment. | Whole bacterial cells |
| Hu et al, 2018 | Whole-Cell *P. aeruginosa* Localized Surface Plasmon Resonance Aptasensor | *P. aeruginosa* | LSPR-based whole-cell sensing with aptamer-based molecular recognition | Utilized a modified Wang et al APT | Diagnostic | Whole bacterial cells |
| Shin et al, 2019 | Detection of Gram-negative bacterial outer membrane vesicles using DNA aptamers. | *E. coli DH5α*<br>*E. coli K12*<br>*Serratia marcescens* | Toggle-cell SELEX (DNA) | Various concentrations of aptamers labelled with FAM were added to 10<sup>8</sup> cells of bacteria. After measuring the fluorescence signal, the binding saturation curves were fitted using non-linear regression model. The dissociation constants (Kd) were calculated against three bacterial strains tested. | Diagnostic<br>GN6<br>GN12 | Whole bacterial cells |
| Roushani M, Sarabaegi M and Pourahmad F, 2019. | Impedimetric aptasensor for *P. aeruginosa* by using a glassy carbon electrode modified with silver nanoparticles. | *P. aeruginosa* | SELEX (DNA) | Utilized a modified Wang et al APT | Diagnostic | Whole bacterial cells |
| Shi X, Zhang J and He F | A new aptamer/polyadenylated DNA interdigitated gold electrode piezoelectric sensor for rapid detection of *P. aeruginosa*. | *P. aeruginosa* | SELEX (DNA) | Utilized a modified Wang et al APT | Diagnostic | Whole bacterial cells |
| Zhao et al, 2019. | C4-HSL aptamers for blocking quorum sensing and inhibiting biofilm formation in *P. aeruginosa* and its structure prediction and analysis. | *P. aeruginosa* | SELEX (DNA) | The assembled Bead-C50-Pool99-SQ was incubated with C4-HSL at different concentrations for 1 hour and the fluorescence intensity was measured, and the aptamer dissociation was calculated, as well as the dissociation constant (Kd). | Inhibit the formation of biofilms of *P. aeruginosa* | Against C4-HSL produced by the *rhl* system. |
| Chen et al, 2019 | A novel method combining aptamer-Ag10NPs based microfluidic biochip with bright field imaging for detection of KPC-2-expressing bacteria. | KPC-2<br>KPC-2-expressing *E. coli* | Protein SELEX and Bacterium SELEX (DNA) | FAM-labelled XK-10 or Sgc was incubated with KPC-2 *E. coli* in different concentrations and measured by flow cytometry. The dissociation constant (Kd) was calculated. | Diagnostic KPC-2 producers<br>XK10 | KPC-2 was used as target protein and KPC-2 *E. coli* was used as target bacterium. |
| Yu et al, 2020 | Screening aptamers for serine β-lactamase-expressing bacteria with Precision-SELEX | KPC-2<br>KPC-2-expressing *E. coli* | Protein SELEX and Bacterium SELEX (DNA) | Utilized (XK10) Chen et al APT | Diagnostic KPC-2 producers<br>XK10 | KPC-2 was used as target protein and KPC-2 *E. coli* was used as target bacterium. |
| Kubiczek et al, 2020 | The diversity of a polyclonal FluCell-SELEX library outperforms individual aptamers as emerging diagnostic tools for identification of carbapenem resistant *P. aeruginosa*. | *P. aeruginosa* | FluCell-SELEX (DNA) | The library outperformed single aptamers targeting different clinically relevant strains, based affinity purification and mass spectrometry. The combined analysis of affinity and specificity evolved confocal laser scanning microscopy (CLSM) and fluorimetry. | Diagnostic | Whole bacterial cells |
| Yang et al, 2020 | A fluorometric assay for rapid enrichment and determination of bacteria by using zirconium-metal organic frameworks as both capture surface and signal amplification tag. | *A. baumannii* | Non-SELEX (DNA) | Utilized a modified Rosoulinjad et al APT | Diagnostic | Whole bacterial cells |
| Yao et al, 2020 | SiC-functionalized fluorescent aptasensor for determination of *Proteus mirabilis*. | *Proteus mirabilis* | Non-SELEX (DNA) | Utilized a modified Savory et al APT | Diagnostic | Whole bacterial cells |
| Su et al, 2020 | Dual aptamer assay for detection of *A. baumannii* on an electromagnetically driven microfluidic platform. | *A. baumannii* | Non-SELEX (DNA) | Utilized a modified Wu et al APT | Diagnostic | Whole bacterial cells |
| Bahari et al, 2020 | Ratiometric fluorescence resonance energy transfer aptasensor for highly sensitive and selective detection of *A. baumannii* bacteria in urine sample using carbon dots as optical nanoprobes | *A. baumannii* | Non-SELEX (DNA) | Utilized a modified Wu et al APT | Diagnostic | Whole bacterial cells |
| Lee et al, 2020 | Electrical antimicrobial susceptibility testing based on aptamer-functionalized capacitance sensor array for clinical isolates. | *A. baumannii*<br>*P. aeruginosa* | Non-SELEX (DNA) | Utilized a modified Yoo et al and Rasoulinejad et al APT | Diagnostic and Therapeutic | Whole bacterial cells |
| Soundy J and Day D, 2020 | Delivery of antibacterial silver nanoclusters to *P. aeruginosa* using species-specific DNA aptamers. | *P. aeruginosa* | Non-SELEX (DNA) | Utilized a modified Soundy and Day APT | Therapeutic | Against live biofilm derived from PA692 *P. aeruginosa* |
| Radhika and Gorthi, 2020 | dsDNA-templated fluorescent copper nanoparticles for the detection of lipopolysaccharides | LPS | Non-SELEX (DNA) | Utilized a modified Kim et al APT | Diagnostic | LPS |
| Krämer M et al, 2021 | BSA Hydrogel Beads Functionalized with a Specific Aptamer Library for Capturing *P. aeruginosa* in Serum and Blood | *P. aeruginosa* | FluCell-SELEX (DNA) | To evaluate specificity the aptamer was incubated in different concentrations with *P. aeruginosa* and others Gram-negative and Gram-positive bacteria, as well as *Candida auris* and submitted to fluorescence microscopy. Then, the fluorescence microscopy images were analyzed using BSFA. | Diagnostic | Whole bacterial cells |

## Results and Discussion

After filter application, 30 papers were selected: 18 regarding non-fermenters (*P. aeruginosa* or *A. baumannii*), seven being aptamers approaching multi species bacteria, and only five regarding a single enterobacteria. Twenty-three papers have used the bacteria whole cell as a target performing *Cell-SELEX* for aptamer selection. The remaining studies performed *Protein-SELEX*. Most of the published papers are regarding aptamers for diagnostic and only seven focus on aptamers for therapeutics. Out of the papers that attempted to use aptamers for therapeutics, none presented successful results. Here, we are going to discuss chronologically the discoveries and development of techniques using aptamers or the SELEX protocol for gram negative bacteria identification, followed by their implementation for diagnostic or therapeutics over the past two decades (Figure 3).

**Figure 3:**
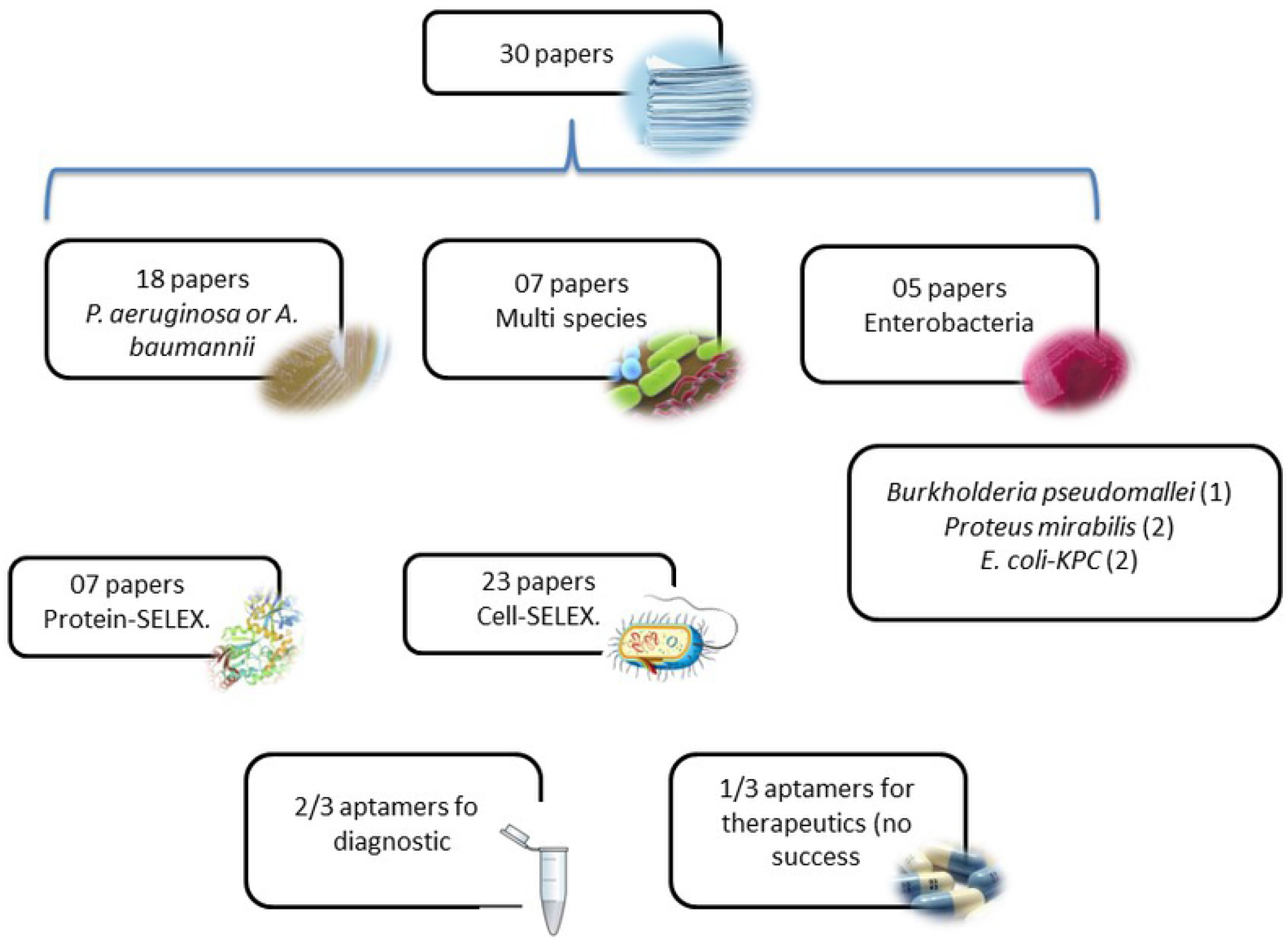
Results graphic summary

Back in 1996, Ocheser and Vasil [9] identified 16 genes regulated by ferric uptake regulator (*Fur*) on *P. aeruginosa* cells using the SELEX technology, which was a pioneer research for the creation of an anti-fur antibody, that through the receptor blockage would reduce the viable iron for the bacterial cell, thus leading to death.

After almost a decade with little progress on aptamer research for gram-negative bacteria, in 2008, Gnamun et al [10], developed a high affinity RNA aptamer for protein recognition from the lethal gram-negative bacteria *Burkholderia pseudomallei* which could be a useful tool for developing a rapid diagnostic assay. However, the *in vitro* selection of aptamers only proceeded until the third round and no final aptamer sequences were obtained from this analysis.

With a distinct approach, Ding et al, 2009 [11], developed an ssDNA aptamer directed against the lipopolysaccharide (LPS) of Gram-negative bacteria, which would provide a broad protection to various agents at once by binding and neutralizing this endotoxin. Even without previous satisfactory results, they achieved valuable findings, such as the importance of the sequence length and composition in the biding and inhibition capacity. Three years later, Kim et al, 2012 [12], also selected aptamers specifically to LPS, using as a target synthesized LPS from *E. coli* 055:B5 Sigma-Aldrich (USA). Using LPS-specific aptamers, they constructed an impedance biosensor on a gold surface that detected LPS in low concentrations, although it showed cross-interaction with other proteins. Still having good potential for practical applications to biological fluids, the LPS-aptamer described by Kim et al was later commercialized by Sigma-Aldrich (USA) and modified for building biosensors for LPS detection [13,14].

To establish a rapid and simple fluorescence in situ hybridization (FISH) method, Wang et al, 2010 [15], selected ssDNA aptamers that bonded specifically to *P. aeruginosa*, using the whole bacteria cell as a target, being the first application of aptamer-FISH for bacterial identification. Successfully, the two selected aptamers showed no cross-reactivity with other bacteria genus, being the first specific aptamer described to *P. aeruginosa*. The initial application of this aptamer was by Yoo et al, 2015 [16], who utilized previously established aptamers to *P. aeruginosa* [15], *L. acidophilus* [8], and *Salmonella typhimurium* [17], to develop a localized surface plasmon resonance (LSPR)-based sensor, the first multiplex mode of bacterial detection using aptamers and capable of detecting low concentrations (30 *cfu)* of three different bacteria on the same assay, in a microchip format.

Savory et al, 2013 [18], developed an in silico maturation (ISM) approach to improve the specificity of aptamers to *Proteus mirabilis* early diagnostic. They applied ISM on five aptamers initially selected by cell-SELEX, increasing the affinity in 36%, leading to the conclusion that the base guanine plays a key role in the aptamer binding capacity across the whole bacteria cell. Later, using this previous described aptamer for *P. mirabilis* [19], Yao et al, 2020 [20], developed a new fluorescent semiconductor nanocrystal for the detection of *P. mirabilis* in blood, urine, and milk. The aptasensor proved to be stable and easily synthesized when tested on milk and simulated forensic blood and urine samples. However, it still needs to be tested in clinical samples with different pathogens concentration.

In 2016, Rasoulinejad and Gargari [21] introduced the first aptamer targeting *A. baumannii* as part of a hybrid detection system utilizing two specific binding molecules (an aptamer (Aci49) and a nanobody against a biofilm associated protein (anti-Bap)), based on a sandwich enzyme linked aptamer sorbent assay (ELASA) system. Which revealed to be promising for rapid detection of *A. baumannii* from clinical isolates, with a low detection threshold (10^3^ CFU). In 2020, still searching for diagnosis of bloodstream infection in early stage, [22] modified, through phosphorylation (p), the previously described aptamers targeting *A. baumanni* (Ab) and LPS [21,23], building a fluorometric assay. The strategy demonstrated an excellent performance to determine *A. baumannii* in blood samples. However, was not able to detect multiple pathogens simultaneously.

Schulmeyer et al, 2016 [24], introduced another application for SELEX on *P. aeruginosa* using SELEX to understand the RNA-binding properties of RsmF which, along with RsmA, is part of the CsrA protein family. They observed that RsmF binding requires target RNAs with two consensus-binding sites (GGA), while RsmA recognizes targets with just a single binding site, both preferring to bind to hexaloop sequences. This data was valuable to define *P. aeruginosa* physiology and virulence.

In 2017, using whole-cell SELEX approach, Soundy and Day [25] identified the first DNA-aptamer highly specific against live biofilm derived from *P. aeruginosa*. Initially, their intention were aptamers with bacteriostatic and/or bactericidal activity; however, even being highly specific, their aptamers did not present therapeutic properties. Three years later, Soundy et al, 2020 [26], designed a promising targeted delivery of silver by minimizing the silver toxicity to the infected host and maximizing the antimicrobial activity. They utilized aptamers previously described by the same group [27] and conjugated to an AgNC scaffolding sequences, being the first described *P. aeruginosa* specific DNA aptamer to deliver a toxic dose of metallic silver as a targeted antimicrobial to *P. aeruginosa* with successful results in invertebrate animals’ model of infection (*Galleria mellonella*).

Looking for aptamers able to target bacteria from different genera, Song et al, 2017 [28], described an *in vitro* isolation method named sequential toggle cell-SELEX (STC-SELEX), where they sought for aptamers sharing the same epitopes. They incubated bacteria from different genera sequentially: *Escherichia coli, Enterobacter aerogenes, Klebsiella pneumoniae, Citrobacter freundii, Bacillus subtilis*, and *Staphylococcus epidermidis*, respectively. After 18 rounds of selection, they arrived at two conserved sequences with broad affinity to the six bacterial genera, as well with a lack of binding affinity to nine other species designated for negative selection. This methodology is potentially useful to find broadly reactive aptamers with affinity to multiple microbiologic genera.

Also looking for broad spectrum aptamers, Graziani et al, 2017 [29], described two aptamers targeting the outer cell wall peptidoglycan, presenting binding affinity to seven different species of Gram-negative and four Gram-positive bacteria. The highest binding activity was found in *K. pneumoniae, P. aeruginosa*, and *A. baumannii* for both aptamers, which probably reflects differences in the cell wall peptidoglycan found in these strains. Broadly specific aptamers with capability to detect a variety of bacterial agents of sepsis could provide a culture- and amplification-free instrument for sepsis diagnostic.

As an alternative for traditional antibody-based assays, Wu et al, 2018 [30], developed an electromagnetically driven microfluidic device which achieved bacterial detection via a nitrocellulose membrane-based dual-aptamer assay. The declared aptamer was selected though SELEX technology and was highly specific against *A. baumannii*. This microfluidic technology was hypothesized to be capable of replacing the conventional antibody assay, forming a sandwich-like structure able to detect the bacteria in about 40 min, being a promising point-of-care bacterial diagnostics instrument. In order to reduce time-consuming diagnostic tools, Su et al, 2020 [31], amino modified the *A. baumannii* aptamer described by Wu et al, 2018 [32], and built a novel dual-aptamer assay [33], a microfluidic device where aptamers were conjugated with magnetic beads to capture the bacteria and with quantum dots to quantify the amount of bacteria. This fully automated system might be promising in remote locations. With a similar objective, Bahari et al, 2020 [34], also utilized Wu et al’s, 2018, aptamer to develop a fluorescent biosensor based on fluorescence resonance energy transfer with ortho-phenylenediamines carbon and nitrogen-doped carbon nanodots as donor’s species and graphene oxide as acceptor for detection of *A. baumannii*. The fluorescent aptasensor can detect bacteria in low concentrations and presents promising results for detection of *A. baumannii* in urine samples.

As a new strategy for biofilm control, Wang et al, 2018 [35], conjugated their previously described aptamer against *P. aeruginosa* (PA-ap1) [15] with cipro?oxacin (CPX), enhancing the efficiency of antibiofilm agents. *P. aeruginosa* was considered as a model organism due to its strong biofilm formation ability and for developing high resistance, as many drugs cannot diffuse into the biofilm. They constructed complexes using PA-ap1, CPX, and single-walled carbon nanotubes (SWNTs) in different combinations. The in vitro results suggest that the aptamer-CPX-SWNTs complex may be a useful approach to control biofilm formation by a variety of bacteria, just changing the aptamer sequence and the target.

Several groups have been working on rapid diagnostic and treatment for Gram-negative agents. Not differently, Hu et al, 2018 [36], used localized surface plasmon resonance (LSPR), a label-free detection system which is non-specific to any type of molecular target. Additionally, a key property of LSPR-based biosensors is the relatively small sensing volume, typically limited to 10-100nm of the surface. To address this problem, a LSPR based sensing platform was developed to detect the whole *P. aeruginosa* cell using a surface-confined aptamer (PA-ap1) previously discovered by Wang et al, 2011 [15]. Applying this approach, *P. aeruginosa* cells are directly pulled down from the solution onto a sensor surface by a surface-confined aptamer, which then does the cell-recognition. Overall, this LSPR sensing platform is characterized by rapid detection and high sensitivity, representing an important advancement in pathogen diagnostics. The LSPR platform supports broader implementation for different pathogens. To prove the method selectivity and versatility, one year later, the same group used the technology to specifically detect *A. baumannii* cells over other Gram-negative (*P. aeruginosa, E. coli*) and Gram-positive (*B. cereus*) bacteria, using a siderophore-Fe(III) complex [37] as an affinity reagent.

Likewise, Roushani et al, 2019 [38], using the *P. aeruginosa* specific aptamer [39], amino modified by Zhong et al, 2018 [40], developed a selective aptasensor on the glassy carbon electrode surface for *P. aeruginosa* ultrasensitive detection. The aptamer amino group binds covalently with AgNPs electrodeposited on the glassy surface, keeping the aptamer attached to the surface. This was the first impedimetric aptasensor used for diagnosis of *P. aeruginosa* in serum samples. Consequently, this procedure could facilitate clinical diagnosis of *P. aeruginosa*.

Also using Wang et al [15] previously described aptamer, Shi et al, 2019 [41], developed a novel sensing method, utilizing magnetic bead-aptamer-Poly(A) DNA complex coupled with an Au multi-channel piezoelectric quartz crystal technology (IDE-MSPQC) for rapid and selective detection of *P. aeruginosa* in sterile defibrinated sheep blood samples. In a way that the Poly-A DNA was complementary to the aptamer and the magnetic bead was employed to immobilize the *P. aeruginosa* aptamer in a sandwich-type structure. This strategy provided a new path for the detection of *P. aeruginosa*; however, it was not detected from the real patients’ samples.

Regardless of the extensive research for bacterial detection methods, none fully meet the demands for real life situations, such as detecting living bacteria in blood. And trying to solve this problem, Shin et al, 2019 [42], isolated two single-stranded DNA aptamers (GN6 and GN12) against *Serratia marcescens, Escherichia coli DH5α*, and *E. coli K12* using the previously described Toggle-cell-SELEX [28]. The bacteria cells were sequentially incubated with a pool of random DNA, selecting aptamers with strong binding affinity to the three selected agents. Using the GN6 aptamer, they assembled an aptamer-based detection tool, an Enzyme-linked aptamer assay (ELAA) to detect bacterial outer membrane vesicles (OMVs) of Gram-negative bacteria, which contain several outer membrane proteins with potent immunostimulatory effects. The GN6-ELAA showed great potential to facilitate medical diagnosis, based on the ability to identify multiple Gram-negative species. Regarding cross-reactivity, a binding assay was performed using Gram-negative and Gram-positive bacterial cells. The aptamers showed high binding affinity to the Gram-negative and almost none to the Gram-positive, suggesting that the unknown targets are exclusively expressed in Gram-negative bacteria.

Besides the diagnostic concerns, *P. aeruginosa* treatment was also a constant challenge and urging for innovative technologies. Pseudomonas aeruginosa biofilm, for example, is extremely difficult to be penetrated by the antibiotics as most of the bacteria are encapsulated in the polysaccharide matrix. It is well known that the rhl and las system affect the biofilm formation and its maintenance, Zhao et al [43] used SELEX protocol to select aptamers with high affinity against the signal molecule C4-HSL secreted by the bacteria, which is part of the rhl system. Theoretically, by depressing the rhl system, the C4-HSL would be held in low level and the biofilm formation would be inhibited. The aptamers selected in this study can reduce the biofilm formation but caused no effect on the normal proliferation of bacteria. Based on these results, these aptamers still could be optimized and used for the development of drugs and detection reagents.

In a continuous effort to potentially aid antimicrobial stewardship, Lee et al, 2020 [44], took over previously descried specific aptamers for six different pathogenic bacteria, and developed a rapid antimicrobial susceptibility test (e-AST). These aptamers included the previously mentioned aptamers for *P. aeruginosa* and *A. baumanii* [45,46]. The algorithm screens bacterial behaviors in real time, thereby allowing the determination of antimicrobial susceptibility within six hours. Even with a higher cost when compared with gold standard broth microdilution (BMD), it can be a lifesaving tool once the discrepancies between e-AST and BMD tests were estimated to be lower than 2.20% when applied on six clinical strains from septic patients.

Kubiczec et al, 2020 [47], introduced the concept of polyclonal aptamer library as a new tool to fight against threating MDR pathogens. Utilizing a variation of the SELEX process through monitoring the successive selection rounds by fluorescence measurements of the developing libraries, a technique called FullCell-SELEX, modified from the earlier described FluMag-SELEX [48] was developed?. They utilized a synthesized library with 40 different aptamer sequences and performed 16 SELEX cycles. They observed a rise of binding affinity throughout the cycles, which suggested a compensation from the larger available aptamer sequences to other targets on the cell surfaces. However, the bacteria recovered motility between day one and three of the experiment. One year later, Kramer et al, 2021 [49], used a focused library from FLuCell-SELEX with eight aptamers against *P. aeruginosa*, which were better suited to consistently bind carbapenem-resistant *P. aeruginosa*. They developed a BSA beads coated anti-*P. aeruginosa* aptamers, which successfully captured cells of this pathogen in human fluids.

So far, none of the studies regarding aptamers for Gram-negative bacteria have approached specifically antibiotic resistant bacteria. Chen et al, 2020 and Yu et al, 2021 [50,51], selected aptamers specifically for KPC-2 producing bacteria (XK10) in a method called Precision-SELEX. In this method, they combined protein SELEX and bacterium SELEX running first four rounds of protein SELEX using KPC-2 as target and subsequently four rounds of conventional bacteria SELEX using *E. coli* KPC-2 producer. Afterwards, they developed a micro-fluid bioship integrated with silver modified aptamers (Biotin-Ag-XK10), which can detect the whole bacteria cell from fluids, giving simplicity in bacterial recognition and detection. The system is promising because it’s able to detect and even differentiate types of carbapenemases in one hour.

Although the number of publications regarding the development of aptamers for pathogenic enterobacteria and non-fermenters has increased over the years, much remains to be done. Only nine of the 30 publications reviewed are regarding the detection of new aptamers for a specific target. The past three years of publications mostly focus on the manufacturing of diagnostic devices using previously described aptamers, without the development of new aptamers. In addition, the publications that approach the use of aptamers as a therapy for gram-negative bacteria have not shown promising results. Another trend in aptamer research involving Gram-negative bacteria is the shift from protein-SELEX to whole-cell-SELEX over the years, and more recently with the combination of both with the so-called Precision-SELEX.

## Conclusion

Even while approaching a potentially life-saving technology, the studies regarding aptamers for Gram-negative bacteria are still limited. The number of aptamers with strong and specific binding capacity are reduced and the challenges remain in eliminating cross-binding activities to other molecules in many real-world complex samples, such as biological fluids. Improving the current selection arrangement (SELEX) and developing more aptamers remains the major barrier for aptamer-related studies. A carefully designed selection preparation can significantly improve the specificity of the identified oligonucleotides, which can better distinguish closely related molecules at low concentrations. As the field of ssDNA-based biosensors continues to grow, improvements in the methodology will be necessary to more rapidly isolate aptamers with the desired affinity and specificity.

## Data Availability

All relevant data are within the manuscript and its Supporting Information files.

## References

1. Delcour AH. Outer membrane permeability and antibiotic resistance. Biochim Biophys Acta -Proteins Proteomics [Internet]. 2009;1794:808–16. Available from: https://linkinghub.elsevier.com/retrieve/pii/S1570963908003592

2. Tuerk C, Gold L. Systematic evolution of ligands by exponential enrichment: RNA ligands to bacteriophage T4 DNA polymerase. Science (80-). 1990;249:505–10.

3. Kimoto M, Yamashige R, Matsunaga K, Yokoyama S, Hirao I. Generation of high-affinity DNA aptamers using an expanded genetic alphabet. Nat Biotechnol. 2013;31:453–7.

4. Bruno JG, Kiel JL. In vitro selection of DNA aptamers to anthrax spores with electrochemiluminescence detection. Biosens Bioelectron [Internet]. 1999;14:457–64. Available from: https://linkinghub.elsevier.com/retrieve/pii/S0956566399000287

5. Tuerk C, Gold L. Systematic Evolution of Ligands by Exponential Enrichment: RNA Ligands to Bacteriophage T4 DNA Polymerase [Internet]. 2007. Available from: www.sciencemag.org

6. Ellington AD, Szostak JW. In vitro selection of RNA molecules that bind specific ligands. Nature [Internet]. 1990;346:818–22. Available from: http://www.nature.com/articles/346818a0

7. Chen F, Zhou J, Luo F, Mohammed A-B, Zhang X-L. Aptamer from whole-bacterium SELEX as new therapeutic reagent against virulent Mycobacterium tuberculosis. Biochem Biophys Res Commun [Internet]. 2007;357:743–8. Available from: https://linkinghub.elsevier.com/retrieve/pii/S0006291X07007097

8. Hamula CLA, Zhang H, Guan LL, Li X-F, Le XC. Selection of aptamers against live bacterial cells. Anal Chem [Internet]. 2008 [cited 2019 Apr 20];80:7812–9. Available from: https://pubs.acs.org/doi/10.1021/ac801272s

9. Ochsner UA, Vasil ML. Gene repression by the ferric uptake regulator in Pseudomonas aeruginosa: cycle selection of iron-regulated genes. Proc Natl Acad Sci U S A [Internet]. 1996 [cited 2019 Apr 20];93:4409–14. Available from: http://www.ncbi.nlm.nih.gov/pubmed/8633080

10. Gnanam AJ, Hall B, Shen X, Piasecki S, Vernados A, Galyov EE, et al. Development of aptamers specific for potential diagnostic targets in Burkholderia pseudomallei. Trans R Soc Trop Med Hyg [Internet]. 2008 [cited 2019 Apr 20];102 Suppl 1:S55-7. Available from: https://academic.oup.com/trstmh/article-lookup/doi/10.1016/S0035-9203(08)70015-4

11. Ding JL, Gan ST, Ho B. Single-stranded DNA oligoaptamers: Molecular recognition and LPS antagonism are length-and secondary structure-dependent. J Innate Immun. 2008;

12. Kim S-EE, Su W, Cho M, Lee Y, Choe W-SS. Harnessing aptamers for electrochemical detection of endotoxin. Anal Biochem [Internet]. Elsevier Inc.; 2012;424:12–20. Available from: https://linkinghub.elsevier.com/retrieve/pii/S0003269712001066

13. Su W, Kim SE, Cho M, Nam J Do, Choe WS, Lee Y. Selective detection of endotoxin using an impedance aptasensor with electrochemically deposited gold nanoparticles. Innate Immun. 2013. p. 388–97.

14. N. K. RGorthi SS. DsDNA-templated fluorescent copper nanoparticles for the detection of lipopolysaccharides. Anal Methods. Royal Society of Chemistry; 2021;13:186–91.

15. Wang K-Y, Zeng Y-L, Yang X-Y, Li W-B, Lan X-P. Utility of aptamer-fluorescence in situ hybridization for rapid detection of Pseudomonas aeruginosa. Eur J Clin Microbiol Infect Dis [Internet]. 2011 [cited 2019 Apr 16];30:273–8. Available from: http://link.springer.com/10.1007/s10096-010-1074-0

16. Yoo SM, Kim D-K, Lee SY. Aptamer-functionalized localized surface plasmon resonance sensor for the multiplexed detection of different bacterial species. Talanta [Internet]. 2015 [cited 2019 Apr 16];132:112–7. Available from: https://linkinghub.elsevier.com/retrieve/pii/S0039914014007619

17. Joshi R, Janagama H, Dwivedi HP, Senthil Kumar TMA, Jaykus L-A, Schefers J, et al. Selection, characterization, and application of DNA aptamers for the capture and detection of Salmonella enterica serovars. Mol Cell Probes [Internet]. 2009 [cited 2019 Apr 20];23:20–8. Available from: https://linkinghub.elsevier.com/retrieve/pii/S0890850808000649

18. Savory N, Lednor D, Tsukakoshi K, Abe K, Yoshida W, Ferri S, et al. In silico maturation of binding-specificity of DNA aptamers against Proteus mirabilis. Biotechnol Bioeng. 2013;

19. Savory N, Lednor D, Tsukakoshi K, Abe K, Yoshida W, Ferri S, et al. In silico maturation of binding-specificity of DNA aptamers against Proteus mirabilis. Biotechnol Bioeng [Internet]. 2013 [cited 2019 Apr 20];110:2573–80. Available from: http://doi.wiley.com/10.1002/bit.24922

20. Yao W, Shi J, Ling J, Guo Y, Ding C, Ding Y. SiC-functionalized fluorescent aptasensor for determination of Proteus mirabilis. Microchim Acta. Microchimica Acta; 2020;187:1–8.

21. Rasoulinejad S, Gargari SLM. Aptamer-nanobody based ELASA for specific detection of Acinetobacter baumannii isolates. J Biotechnol [Internet]. 2016 [cited 2019 Apr 16];231:46–54. Available from: https://linkinghub.elsevier.com/retrieve/pii/S0168165616302802

22. Yang S, Guo Y, Fan J, Yang Y, Zuo C, Bai S, et al. A fluorometric assay for rapid enrichment and determination of bacteria by using zirconium-metal organic frameworks as both capture surface and signal amplification tag. Microchim Acta. Microchimica Acta; 2020;187.

23. Wang N, Dai H, Sai L, Ma H, Lin M. Copper ion-assisted gold nanoparticle aggregates for electrochemical signal amplification of lipopolysaccharide sensing. Biosens Bioelectron. Elsevier; 2019;126:529–34.

24. Schulmeyer KH, Diaz MR, Bair TB, Sanders W, Gode CJ, Laederach A, et al. Primary and Secondary Sequence Structure Requirements for Recognition and Discrimination of Target RNAs by Pseudomonas aeruginosa RsmA and RsmF. J Bacteriol. 2016;

25. Soundy J, Day D. Selection of DNA aptamers specific for live Pseudomonas aeruginosa. Omri A, editor. PLoS One [Internet]. 2017 [cited 2019 Apr 16];12:e0185385. Available from: https://dx.plos.org/10.1371/journal.pone.0185385

26. Soundy J, Day D. Delivery of antibacterial silver nanoclusters to Pseudomonas aeruginosa using species-specific DNA aptamers. J Med Microbiol. 2020;69:640–52.

27. Soundy J, Day D. Selection of DNA aptamers specific for live Pseudomonas aeruginosa. Omri A, editor. PLoS One [Internet]. 2017 [cited 2019 Apr 16];12:e0185385. Available from: https://dx.plos.org/10.1371/journal.pone.0185385

28. Song MY, Nguyen D, Hong SW, Kim BC. Broadly reactive aptamers targeting bacteria belonging to different genera using a sequential toggle cell-SELEX. Sci Rep [Internet]. 2017 [cited 2019 Apr 16];7:43641. Available from: http://www.nature.com/articles/srep43641

29. Graziani AC, Stets MI, Lopes ALK, Schluga PHC, Marton S, Ferreira IM, et al. High efficiency binding aptamers for a wide range of bacterial sepsis agents. J Microbiol Biotechnol. 2017;

30. Wu J-H, Wang C-H, Ma Y-D, Lee G-B. A nitrocellulose membrane-based integrated microfluidic system for bacterial detection utilizing magnetic-composite membrane microdevices and bacteria-specific aptamers. Lab Chip [Internet]. 2018 [cited 2019 Apr 16];18:1633–40. Available from: http://xlink.rsc.org/?DOI=C8LC00251G

31. Su CH, Tsai MH, Lin CY, Ma YD, Wang CH, Chung Y Da, et al. Dual aptamer assay for detection of Acinetobacter baumannii on an electromagnetically-driven microfluidic platform. Biosens Bioelectron [Internet]. Elsevier B.V.; 2020;159:112148. Available from: https://doi.org/10.1016/j.bios.2020.112148

32. Wu JH, Wang CH, Ma YD, Lee G Bin. A nitrocellulose membrane-based integrated microfluidic system for bacterial detection utilizing magnetic-composite membrane microdevices and bacteria-specific aptamers. Lab Chip. 2018;

33. Wang CH, Wu JJ, Lee G Bin. Screening of highly-specific aptamers and their applications in paper-based microfluidic chips for rapid diagnosis of multiple bacteria. Sensors Actuators, B Chem. 2019;

34. Bahari D, Babamiri B, Salimi A, Salimizand H. Ratiometric fluorescence resonance energy transfer aptasensor for highly sensitive and selective detection of Acinetobacter baumannii bacteria in urine sample using carbon dots as optical nanoprobes. Talanta [Internet]. Elsevier B.V.; 2021;221:121619. Available from: https://doi.org/10.1016/j.talanta.2020.121619

35. Wang S, Mao B, Wu M, Liang J, Deng L. Influence of aptamer-targeted antibiofilm agents for treatment of Pseudomonas aeruginosa biofilms. Antonie van Leeuwenhoek, Int J Gen Mol Microbiol. 2018;

36. Hu J, Fu K, Bohn PW. Whole-Cell Pseudomonas aeruginosa Localized Surface Plasmon Resonance Aptasensor. Anal Chem [Internet]. 2018 [cited 2019 Apr 16];90:2326–32. Available from: http://pubs.acs.org/doi/10.1021/acs.analchem.7b04800

37. Hu J, Ghosh M, Miller MJ, Bohn PW. Whole-cell biosensing by siderophore-based molecular recognition and localized surface plasmon resonance. Anal Methods. 2019;

38. Roushani M, Sarabaegi M, Pourahmad F. Impedimetric aptasensor for Pseudomonas aeruginosa by using a glassy carbon electrode modified with silver nanoparticles. Microchim Acta. Microchimica Acta; 2019;186.

39. Wang K-Y, Zeng Y-L, Yang X-Y, Li W-B, Lan X-P. Utility of aptamer-fluorescence in situ hybridization for rapid detection of Pseudomonas aeruginosa. Eur J Clin Microbiol Infect Dis [Internet]. 2011 [cited 2019 Apr 20];30:273–8. Available from: http://link.springer.com/10.1007/s10096-010-1074-0

40. Zhong Z, Gao X, Gao R, Jia L. Selective capture and sensitive fluorometric determination of Pseudomonas aeruginosa by using aptamer modified magnetic nanoparticles. Mikrochim Acta [Internet]. 2018 [cited 2019 Apr 16];185:377. Available from: http://link.springer.com/10.1007/s00604-018-2914-3

41. Shi X, Zhang J, He F. A new aptamer/polyadenylated DNA interdigitated gold electrode piezoelectric sensor for rapid detection of Pseudomonas aeruginosa. Biosens Bioelectron [Internet]. Elsevier B.V.; 2019;132:224–9. Available from: https://doi.org/10.1016/j.bios.2019.02.053

42. Shin HS, Gedi V, Kim JK, Lee D ki. Detection of Gram-negative bacterial outer membrane vesicles using DNA aptamers. Sci Rep [Internet]. Springer US; 2019;9:1–8. Available from: http://dx.doi.org/10.1038/s41598-019-49755-0

43. Zhao M, Li W, Liu K, Li H, Lan X. C4-HSL aptamers for blocking qurom sensing and inhibiting biofilm formation in Pseudomonas aeruginosa and its structure prediction and analysis. PLoS One. 2019;14:1–17.

44. Lee KS, Lee SM, Oh J, Park IH, Song JH, Han M, et al. Electrical antimicrobial susceptibility testing based on aptamer-functionalized capacitance sensor array for clinical isolates. Sci Rep [Internet]. Nature Publishing Group UK; 2020;10:1–9. Available from: https://doi.org/10.1038/s41598-020-70459-3

45. Yoo SM, Kim DK, Lee SY. Aptamer-functionalized localized surface plasmon resonance sensor for the multiplexed detection of different bacterial species. Talanta. 2015;

46. Rasoulinejad S, Gargari SLM. Aptamer-nanobody based ELASA for specific detection of Acinetobacter baumannii isolates. J Biotechnol [Internet]. 2016 [cited 2019 Oct 22];231:46–54. Available from: http://www.ncbi.nlm.nih.gov/pubmed/27234880

47. Kubiczek D, Raber H, Bodenberger N, Oswald T, Sahan M, Mayer D, et al. The Diversity of a Polyclonal FluCell-SELEX Library Outperforms Individual Aptamers as Emerging Diagnostic Tools for the Identification of Carbapenem Resistant Pseudomonas aeruginosa. Chem -A Eur J. 2020;26:14536–45.

48. Stoltenburg R, Reinemann C, Strehlitz B. FluMag-SELEX as an advantageous method for DNA aptamer selection. Anal Bioanal Chem. 2005;383:83–91.

49. Krämer M, Kissmann AK, Raber HF, Xing H, Favella P, Müller I, et al. Bsa hydrogel beads functionalized with a specific aptamer library for capturing pseudomonas aeruginosa in serum and blood. Int J Mol Sci. 2021;22.

50. Chen J, Li H, Xie H, Xu D. A novel method combining aptamer-Ag10NPs based microfluidic biochip with bright field imaging for detection of KPC-2-expressing bacteria. Anal Chim Acta [Internet]. 2020 [cited 2020 Oct 8];1132:20–7. Available from: https://linkinghub.elsevier.com/retrieve/pii/S0003267020307996

51. Yu F, Chen J, Wang Z, Yang H, Li H, Jia W, et al. Screening aptamers for serine β-lactamase-expressing bacteria with Precision-SELEX. Talanta [Internet]. Elsevier B.V.; 2021;224:121750. Available from: https://doi.org/10.1016/j.talanta.2020.121750

